# Diagnosing Epilepsy with Normal Interictal EEG Using Dynamic Network Models

**DOI:** 10.1101/2023.08.12.23294018

**Authors:** Patrick Myers, Kristin Gunnarsdottir, Adam Li, Vlad Razskazovskiy, Dale Wyeth, Edmund Wyeth, Alana Tillery, Kareem Zaghloul, Sara Inati, Jennifer Hopp, Babitha Haridas, Jorge Gonzalez-Martinez, Anto Bagíc, Joon-yi Kang, Michael Sperling, Niravkumar Barot, Sridevi V. Sarma, Khalil S. Husari

## Abstract

**Objective:** While scalp EEG is important for diagnosing epilepsy, a single routine EEG is limited in its diagnostic value. Only a small percentage of routine EEGs show interictal epileptiform discharges (IEDs) and overall misdiagnosis rates of epilepsy are 20-30%. We aim to demonstrate how analyzing network properties in EEG recordings can be used to improve the speed and accuracy of epilepsy diagnosis - even in the absence of IEDs.

**Methods:** In this multicenter study, we analyzed routine scalp EEGs from 198 patients with suspected epilepsy and normal initial EEGs. The patients’ diagnoses were later confirmed based on an epilepsy monitoring unit (EMU) admission. About 46% ultimately being diagnosed with epilepsy and 54% with non-epileptic conditions. A logistic regression model was trained using spectral and network-derived EEG features to differentiate between epilepsy and non-epilepsy. The model was trained using 10-fold cross-validation on 70% of the data, which was stratified to include equal numbers of epilepsy and non-epilepsy patients in both training and testing groups. The resulting tool was named EpiScalp.

**Results:** EpiScalp achieved an area under the curve (AUC) of 0.940. The model had an accuracy of 0.904, a sensitivity of 0.835, and a specificity of 0.963 in classifying patients as having epilepsy or not.

**Interpretation:** EpiScalp provides accurate diagnostic aid from a single initial EEG recording, even in more challenging epilepsy cases with normal initial EEGs. This may represent a paradigm shift in epilepsy diagnosis by deriving an objective measure of epilepsy likelihood from previously uninformative EEGs.

## Introduction

Epilepsy is a brain disorder characterized by a predisposition to experiencing recurrent seizures, affecting approximately 60 million people worldwide.^1^ Although 8-10% of the population will experience a seizure during their lifetime, only 2-3% of the individuals develop epilepsy.^1^ Evaluation of patients experiencing suspected seizures includes a thorough clinical history and is usually accompanied by a routine scalp EEG and brain imaging.^2^ After determining whether the episode was an epileptic seizure, the clinician evaluates the risk of subsequent seizures and the need to start anti-seizure medications.^3^

Scalp EEG plays a central role in diagnosing epilepsy and evaluating the risk of subsequent seizures.^2,4,5^ Visual analysis and interpretation remain the gold standard in analyzing EEGs.^6^ Neurologists look for abnormalities including spikes and sharp waves, formally called interictal epileptiform discharges (IEDs), in addition to focal slowing of EEG waveform activity, all of which are known indicators of epileptic tendency.^1,4^ Unfortunately, the sensitivity of scalp EEG in diagnosing epilepsy varies from 29-55%, largely due to the sporadic nature of IEDs.^7–9^ Repeat scalp EEGs may increase the sensitivity up to 92%, but lead to significant use of financial and logistical resources for both patients and the healthcare system.^7^ Additionally, misinterpretation of the EEG as being abnormal^10^ and overinterpretation of EEG are major contributors to misdiagnosis and may result in unnecessary pharmacotherapy and reduce patient quality of life.^8^

Visual interpretation of EEG recordings is subjective and prone to variability across different EEG readers.^9,11–13^ As a result, both false positive and false negative diagnoses commonly occur and overall misdiagnosis rates of epilepsy are nearly 30%.14,15

Computational EEG analysis tools have emerged recently to assist clinicians in EEG analysis.^16^ The majority of the proposed algorithms automate detection of abnormalities, and a widely used commercially available software is Persyst’s Spike Detector,^13,17^ which identifies possible spikes in the EEG recording. There have been several promising publications analyzing EEGs in search of features to distinguish epilepsy from EEGs void of abnormalities. Many such papers, however, use healthy individuals as their control group which limits their utility as this setup is both not representative of a real-world clinical setting and may not yield an indicator that is specific to epilepsy.^16,18–23^

Two network-based metrics, fragility^24^ and source-sink^25^, have been proposed to aid in the localization of the epileptogenic zone from ictal and interictal intracranial EEG recordings, respectively. Both methods operate on the assumption that there are alterations in the neural network, specifically in the epileptic tissue, of an epilepsy patient.^26–28^ We applied these concepts to scalp EEG to differentiate the network of epileptic and non-epileptic EEGs. We named the tool that combines these analyses *EpiScalp*.

*EpiScalp* aims to diagnose epilepsy from a patient‘s scalp EEG by i) estimating patient-specific dynamical network models from the EEG recording^24,25^ and ii) analyzing the network properties to detect whether pathological patterns, inherent to an epileptic brain, are present in the EEG network.

## Methods

### Subjects

This was a multi-center analysis of retrospectively collected data (**Table 1**). Adults patients admitted to the Epilepsy Monitoring Unit (EMU) were screened to be included in the study. Patients who did not have a habitual event/seizure in the EMU and patients with both epileptic and non-epileptic seizures were excluded. Potential candidates were labeled as having either epileptic seizures or non-epileptic events, based on video EEG results in the EMU. Most patients with non-epileptic events had functional seizures (FS).^29^

Demographic and clinical data were collected from the electronic medical charts.

The first EEG available at the center was collected for this study. Further exclusion criteria included patients without routine EEG available in the center, patients with IED or temporal intermittent rhythmic delta activity (TIRDA) based on the EEG report, and patients with EEG containing continuous, large myogenic artifacts or technical problems. In the course of standard clinical practice, EEGs were reviewed and interpreted by a fellowship trained credentialed neurologist.

**Figure 1** depicts how patients were selected.

### Data

All participating centers record scalp EEG using the standard 10-20 montage scheme. Recording sampling rate varied based on the location: 200 Hz at Johns Hopkins Hospital (JHH) and Johns Hopkins Bayview Medical Center (JHBMC), 256 Hz at University of Pittsburgh Medical Center (UPMC) and University of Maryland Medical Center (UMMC), and either 500 or 1000 Hz at Thomas Jefferson University Hospital (TJUH). All EEG records were downsampled to 200 Hz for analysis. Signals were referenced against an average of the C3 and C4 electrodes, and thus the reference electrodes were excluded from further analysis. Since not all centers recorded from the midline channels (Fz, Pz, and Cz), we opted to discard signals acquired from these contacts. The remaining 14 EEG channels were included for analysis. Data was stored in European Data Format (EDF) and organized using the BIDS-EEG scheme.

### Preprocessing

To remove most myogenic artifacts, a second order bandpass filter between 1 and 30 Hz was applied to each record. The remaining artifacts, mostly ocular and cardiac, were removed through an automated process using Independent Component Analysis (ICA). Preprocessing was performed in Python via the package MNE^30^. The sub-package MNE-ICA was used to automatically calculate the independent components from the filtered signals and MNE-ICLabel was used to classify each component as either EEG signal or one of the following artifact types: eye, muscle, line noise, or other^31^. MNE-ICLabel returns a percent likelihood for each classification. Components with less than 30% probability of containing EEG signal were removed. The remaining components were reconstructed into a cleaned EEG record that was used for the remaining analysis.

### Network-Based EEG Metrics

Two network based metrics, neural fragility^24^ and source-sink index^25^, have been shown to be useful in localizing the epileptogenic zone (EZ) in ictal and interictal stereo EEG (sEEG) recordings, respectively. Both methods were validated across multiple epilepsy centers and developed in the Sarma lab.^24,25,32^

Neural fragility is a concept related to the underlying dynamics of epileptic networks and the emergence of seizures. Specifically, it suggests that the onset of focal seizures may be related to the presence of a few fragile nodes, which render the epileptic network unstable and susceptible to seizure activity. The idea is that during interictal periods, the network is in a “balanced” state, meaning that activity hovers around a baseline value and can respond transiently to perturbations but returns to the baseline value. However, during a pre-ictal period and during a seizure event, the network becomes “unbalanced”, with activity growing in amplitude, oscillating, and spreading throughout the brain. The notion of balance refers to the level of inhibitory and excitatory neuronal populations across the brain network.

The theory of fragility in a dynamical network is presented by Sritharan and Sarma.^33^ Fragility computed from intracranial EEG (iEEG) recordings is described by Li et al^24,28^. Briefly, the fragility of each iEEG channel is computed by first estimating a linear time varying dynamical network model from iEEG data before, during and after a seizure event.^28,34^ This model consists of a sequence of linear time invariant models of the form:

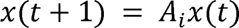

where i=1,2,… and *A_i_* describes how each channel influences each other dynamically within a 500 msec window of the iEEG. Then, from the Ai matrices, an optimization routine is performed to find which channels would cause imbalance with minimal perturbations to their connections to other nodes in the network.

The source-sink index is also derived from the linear time varying dynamical network model, estimated solely from interictal data. The term “source” refers to a group of brain regions that are actively influencing the electrical activity of other regions, while “sink” refers to a group of regions that are mostly being influenced by others’ activity.

In the context of EEG or iEEG data, source-sink connectivity refers to the analysis of how electrical activity propagates through the brain network, from the sources to the sinks. In *Gunnarsdottir et al*, we hypothesize that the epileptogenic zone in a patient is inhibited by other regions during non-clinical seizure periods and thus are sinks.^25^ To investigate this, we created an algorithm that identifies two groups of nodes within the interictal iEEG network. These groups are nodes that continuously inhibit neighboring nodes (’sources’) and the nodes that are inhibited (’sinks’). We estimated patient-specific dynamical network models from several minutes of interictal iEEG data, and the resulting connectivity properties as gleaned from the A matrices helped identify the top sources and sinks within the network. Specifically, we quantified each node using source-sink metrics derived from the A matrices.^25^

In this current study, one fragility metric and three source-sink metrics (the source-sink index, sink-index, and source-influence) were calculated for every EEG recording.

### Spectral Metrics

In addition to the network-based metrics described above, we computed spectral-based metrics from four different frequency bands of interest: Delta (1 – 4 Hz), Theta (4 – 8 Hz), Alpha (8 – 12 Hz), and Beta (12 – 30 Hz). We calculated a multitaper Fourier Transform over non-overlapping 2 second windows to accommodate the slower frequency band. All metrics resulted in an n-by-m matrix where n is the number of channels and m is the number of time windows.

### Dimensionality Reduction

To reduce the dimensionality of the feature space and to remove recording duration as a variable, each metric matrix was summarized into an n-by-1 vector where n is the number of channels. Two summary methods were employed to achieve this dimensionality reduction: a) time-average, and b) Principal Component Analysis (PCA). Since no seizures are present in the EEG recordings (interictal EEG recordings), the signals are relatively stable over time. Thus, we chose to compute the average across time windows for each channel. In contrast, the second summary method aimed to capture some time-varying dynamics through PCA. To maintain the meaning of channels and only reduce dimensions on the time axis, each channel’s time series was projected onto each of the first two principal components to result in a single value per channel.

### Lobe and Channel Feature-Categories

Two main feature categories were derived from each of the two dimension-reduced metric vectors: lobe-based features and channel-based features. Lobe based features were calculated by aggregating channels that belonged to the same lobe and then calculating statistics (such as the mean and standard deviation) within each lobe. Channel based features were calculated by analyzing the quantiles (10%, 50%, 90%, mean, and standard deviation) of the individual channel values over time.

This two-step feature generation method was repeated for every metric (i.e., fragility, the three source-sink, and time-frequency metrics), resulting in 39 features per metric, or 312 unique features.

### Model Design

We modeled the probability of a patient having epilepsy by constructing a logistic regression model using the features described above as covariates. Since the possible feature space was too large for the sample size, feature selection was necessary before constructing a final model. A recursive, greedy feature elimination procedure was developed. First, features were grouped according to their generation method (dimensionality reduction method and feature category), e.g., all PCA lobe-based features or time-averaged channel-based features. Within each such group, initial logistic regression models (described below) were built from all possible combinations of metrics. After this search over feature groups, all resulting models were ranked by their predictive performance. Features that consistently appeared in the top performing models, especially those with generally large and significant feature weights, were selected. This process resulted in 21 candidate features.

From these candidate features, a final model was trained. Results presented below are only on this final model. Figure 2 details this procedure.

For each step of this process, we performed a 10-fold cross validation (CV) procedure. Each fold was randomly split into 70% training data and 30% testing data, where the training set had an even split of epilepsy and non-epilepsy patients. The model’s weights were then tuned for each training set and the performance was evaluated on the corresponding test set. A varying threshold was applied to the model’s output (i.e., the probability of each subject belonging to the epilepsy group), to generate evaluation metrics. Applying this threshold classified the probabilities into a predicted diagnosis, which could be compared to the actual diagnosis of each patient. The metrics used were the area under the curve (AUC) of the resulting ROC curve, and the model’s accuracy, sensitivity, and specificity at the optimal threshold. For the feature selection phase, the models were ranked based on the performance on these test sets. We present these results for the final model below.

We chose to use a logistic regression model to keep the models as simple and interpretable as possible. The feature selection method described above was not optimal because there was no way to assess if the strong features from different selection groups carried the same information. To attempt to prevent this problem, we allowed more features to be selected for the final model than should be allowed based on the sample size. We applied an L1-penalty on the logistic regression models^35^ to prevent overfitting from the excess features.

### Standard Protocol Approvals

This study was approved by the institutional review board (IRB) at each contributing center. The data presented and used for analysis were deidentified.

## Results

A total of 198 subjects were selected from 4 epilepsy centers – JHU (n=90), UPMC (n=62), TJUH (n=27), and UMMC (n=19). The final diagnosis (based on the EMU results) of the subjects was 107 epilepsy and 91 non-epilepsy.

### *EpiScalp* Accurately Predicts Epilepsy Status from Indeterminate EEG Records

From each patient’s single scalp EEG record, we computed several network-based metrics which formed the basis for *EpiScalp*’s features. With this trained model, a probability of belonging to the epilepsy group was determined for each patient. Figure 3A shows this predicted probability of epilepsy where each patient is represented by a single dot. The x-axis denotes the patient’s diagnosis (based on gold standard - EMU evaluation). By applying a threshold to these probabilities, the prediction can be converted to a classification. Probabilities above the threshold are classified as epilepsy and below the threshold are classified as non-epilepsy. With each such threshold, we can assess the model’s performance against the patients’ final diagnosis by computing accuracy, sensitivity, and specificity. The threshold that results in the best accuracy (*a*^*^) is shown with the orange line in Figure 3A. With this optimal threshold, *EpiScalp* achieves an accuracy of 0.904, sensitivity of 0.835, and specificity of 0.963. While *a*\* maximizes prediction accuracy, a lower threshold may be preferable if sensitivity is considered more important than specificity. Figure 3B shows an overview of the model’s performance across a varying threshold, represented by a receiver operating characteristic curve (ROC, blue line). The overall model’s performance can be assessed by the AUC from this ROC curve. Across the 10 folds, the model had an average AUC of 0.940.

When used to assess epilepsy risk, *EpiScalp* can provide great utility. We define an “epilepsy highly unlikely” range as any probability less than 0.32. Patients within this category had a 92% (81/88) chance of not having epilepsy. We also define an “epilepsy highly likely” range as any probability greater than 0.61. Patients within this category had a 95% (76/80) chance of having epilepsy. Patients whose predicted probability falls between these ranges have a medium chance of epilepsy and would therefore be considered indeterminate. If *EpiScalp* is used for all patients that do not lie in the indeterminate category, then it would achieve 93% accuracy, 92% sensitivity, and 95% specificity on 168 of our patients.

### *EpiScalp* Performance is Invariant to Patient Type, Age, Sex, Use of AEDs, Sleep State, and Duration of EEG recordings

To verify the generalizability of *EpiScalp* among different patients, we assessed the model’s performance against various demographic and clinical characteristics including sex, age, whether the patient was taking anti-seizure medications during the recording, the amount of time the patient was asleep during the recording, and the duration of the recording. This analysis is displayed in Figure 5.

To quantify our findings, we performed a Z-test between all pairings of variables within a category. There were no significant differences found.

*EpiScalp* was also insensitive to epilepsy type as shown in Figure 6.

### Feature Importance

The final model had 20 features whose coefficients were statistically significantly different from 0 (displayed in Figure 4). Ten of the 20 were network-based features, out of which 7 were derived from source-sink metrics. Of the 10 remaining spectral features, alpha, beta, and delta band features emerged as significant. The sign of each weight determines whether the feature increases or decreases the likelihood of having epilepsy as the feature increases. For example, the coefficient for “Mean of frontal delta” is 1.01 - the more the average delta band power in the frontal lobes, the probability that the patient has epilepsy modulates up by 101%. Or if the standard deviation of the fragility across EEG electrodes is large, then this modulates the probability that the patient has epilepsy up by a factor of over 200%.

## Discussion

### Current Practice

Currently, epilepsy is clinically diagnosed by gathering evidence rather than through a single biomarker or test.^15^ The primary evidence is usually sourced from the clinical patient interview. Neuro-imaging tests, such as MRI and scalp EEG, are also conducted and visually analyzed. The lack of reliable, specific, and easily identifiable biomarkers on these scans limits their usefulness in diagnosis.

Our analysis suggests that network analysis of scalp EEG along with spectral features together are able to provide diagnostic insight for epilepsy, even when no visual abnormalities are present on the record. Since scalp EEG recordings have sporadic prevalence of these visually identified abnormalities, like IEDs, their usefulness is limited in practice. Our findings support the use of *EpiScalp* in cases where scalp EEG shows normal findings. We have defined three suggested risk score thresholds (epilepsy highly unlikely, epilepsy highly likely, and indeterminate) for the interpretation of *EpiScalp*’s output. With the provided thresholds, *EpiScalp* can provide strong evidence (i.e., 93% accuracy) to support or reject an epilepsy diagnosis in 84% (168 of 198) previously normal studies.

### Generalizability of *EpiScalp*

The algorithm’s performance is agnostic to expected patient variation in age and sex, as well as the number of anti-seizure medications the patient was on, whether the patient was asleep or awake during the recording, and the duration of the recording. For epilepsy patients, EpiScalp’s performance was unaffected by the type of epilepsy.

### Network Based EEG Features

Prior work has demonstrated the utility of analyzing neuroimaging data as a network for providing useful insights into the dynamics of an epileptic brain.^24,25^ By assessing the interactions of brain regions as captured through multiple electrodes, this prior work suggested that you can better localize the epileptogenic region of the brain than by using traditional, single-channel analyses. Since these studies rely on invasive recordings, like electrocorticography (ECoG) and stereo-electroencephalography (sEEG), they could not be directly applied to assist in the diagnosis of epilepsy.

In this study, we expand upon the idea that there is some characteristic network abnormality that is always (even during rest) present in an epileptic brain. Since the goal is to diagnose epilepsy, rather than to localize the epileptogenic zone, this analysis needs to provide the same output independent of the type of epilepsy. This alternative goal, paired with the poor spatial density of scalp EEG as compared to invasive methods, meant that we could not simply apply the same methods as in the prior studies.^24,25^ Instead, we derived features from recently introduced network-based metrics, fragility and source-sink, that would ideally identify all patients with epilepsy instead of a specific type or focus. A combination of these network features were used along with spectral features to predict the diagnosis of these patients.

In our analysis, we identified fragility and sink connectivity as two of the top five significant features within the model. A notable finding was that a greater dissimilarity in fragility between the left and right brain hemispheres was associated with a higher likelihood of epilepsy in patients. This observation aligns with our prior research *Li et al*, which demonstrated that epileptogenic zones (EZ) tend to exhibit increased fragility compared to non-epileptic regions, typically localized within one hemisphere of the brain.^24^

Another noteworthy feature among the top five was the variance in sink connectivity between the two brain hemispheres. Specifically, higher variance (or equivalently standard deviation) in sink connectivity between the left and right brain hemispheres was associated with a lower likelihood of epilepsy in patients. This discovery is in line with our previous studies, which show that EZ regions are characterized as strong sinks that are connected to all brain regions, most notably top sources as outlined in *Gunnarsdottir et al* and not just regions in the same hemisphere.^25^

### Spectral Features

Prior studies have shown that patients with epilepsy have different spectral EEG features when compared to healthy controls.^18,36–38^ We found that alpha, delta, and beta bands features emerged as statistically significant in our final model. Specifically, the higher the delta power in the frontal lobe, the higher the likelihood of epilepsy. The higher the variance of alpha power between left and right frontal lobes, the higher the likelihood of epilepsy. Finally, the higher overall beta power in the EEG snapshot, the lower the likelihood of epilepsy.

Our findings were in line with prior studies where slowing of the posterior dominant rhythm (alpha rhythm), increase in delta power, and decrease in beta power have been noted in patients with epilepsy.^18,36–39^ It is important to emphasize that it is the collective several metrics contributed to the model rather than a single analysis. Additionally, all of the EEGs in our study were read as qualitatively normal.

### Existing Computational Tools

In the realm of EEG analysis, there has been a recent emergence of computational tools designed to aid clinicians. These tools primarily focus on automating the detection of abnormalities. One notable software in this domain is Persyst’s Spike Detector,^13,17^ which has gained popularity as a widely utilized commercial solution. It functions by automatically identifying potential spikes within EEG recordings.

Several publications have explored the analysis of EEGs to uncover hidden features that can differentiate between epilepsy and normal EEGs lacking abnormalities. These studies have relied on healthy individuals as their control group.^18–22^ In contrast, our study utilized subjects with conditions mimicking epilepsy (for example functional seizures), hence representing a better applicability to the real-world clinical scenarios.

One notable study, however, compared EEGs of epilepsy patients to patients suffering from other seizure mimicking disorders (e.g., functional seizures) and achieved 71% accuracy.^23^ Unfortunately, the non-interpretable nature of neural network models, as well as the lower accuracy, hinder the clinical utility of such a tool. Our use of a simple logistic regression model allows for interpretable results from *EpiScalp*.

### Limitations

For this study, we aimed to develop a tool, *EpiScalp*, that could compute the probability of epilepsy diagnosis from previously non-diagnostic EEG information. Since our primary input was inconclusive data, we needed an alternative method of confirming the diagnosis of the subjects in this study. The best way to be confident in our labels retrospectively was to only gather patients who had EMU monitoring with video EEG. As shown in Figure 1, we then selected the most traditionally difficult subset of patients, i.e., those with no abnormalities present in their EEG, for this study. While this approach was necessary for clean study labels, we acknowledge that it introduces patient selection bias.

While there are several conditions that may mimic epilepsy, the most common alternative diagnosis determined by an EMU visit at the participating centers was FS. It is possible that our analysis will not generalize well to other mimics, such as syncope. Since we specifically targeted features that have proven useful in analyzing epileptic networks, we believe our analysis is uniquely identifying epilepsy and would therefore be able to differentiate it from any alternative condition. However, a further study would be needed to validate this claim.

All centers who contributed data to this study are specialized, academic epilepsy centers. These centers are likely to see a higher proportion of difficult-to-diagnose cases referred from external clinics. This bias is especially evident when selecting patients based on extended in-patient stays as the specialized centers are more equipped to handle these visits than other clinical settings. Since *EpiScalp*’s input is the most basic neurophysiological data, i.e., a routine scalp EEG with the standard 10-20 montage, we believe our results would hold in a less specialized setting, but this claim needs to be validated.

Despite these limitations, we have devised network-based evidence to support or refute an epilepsy diagnosis from scalp EEG which was previously considered uninformative. The findings in this study suggest it may be possible to shift the diagnosis of epilepsy to a more test-driven diagnosis. EpiScalp could be used in conjunction with clinical reasoning to provide a more confident, faster diagnosis of epilepsy.

### Future Work

There remains significant work to be done before a complete paradigm shift in the diagnostic process can be achieved. Validating this approach on a more representative, prospective patient population would ensure that the tool is unique to epilepsy. Additionally, testing the tool on data collected from various clinical settings, instead of all specialized academic centers as in this study, could help prove the real-world clinical usability of such a metric.

## Data Availability

Data produced in the present study are not currently available.

## Acknowledgements

We would like to thank Dr. Alexandra Urban, and Dr. Vijayalakshmi Rajasekaran of the University of Pittsburgh Medical Center, Dr. Gregory Bergey of Johns Hopkins Hospital, and the Louis B. Thalheimer Fund for Translational Research for supporting this work.

## Author Contributions

Authors PM, KG, AL, MS, NB, SVS, and KSH conceived the study and performed the majority of the data analysis. Authors VR, DW, EW, AT, and BH also assisted with data acquisition and analysis. Authors KZ, SI, JH, JGM, AB, and JK helped design the study and draft the manuscript.

---

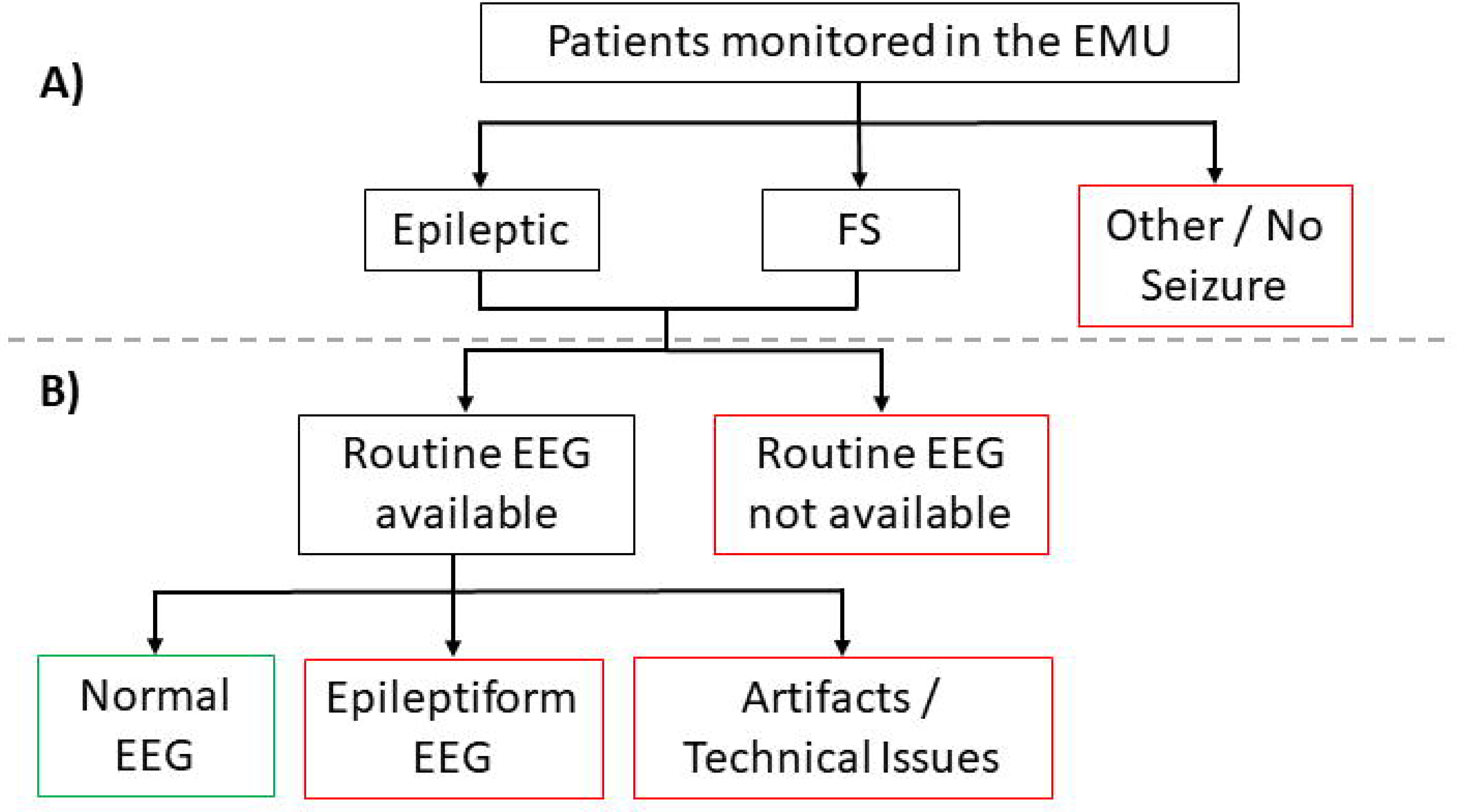

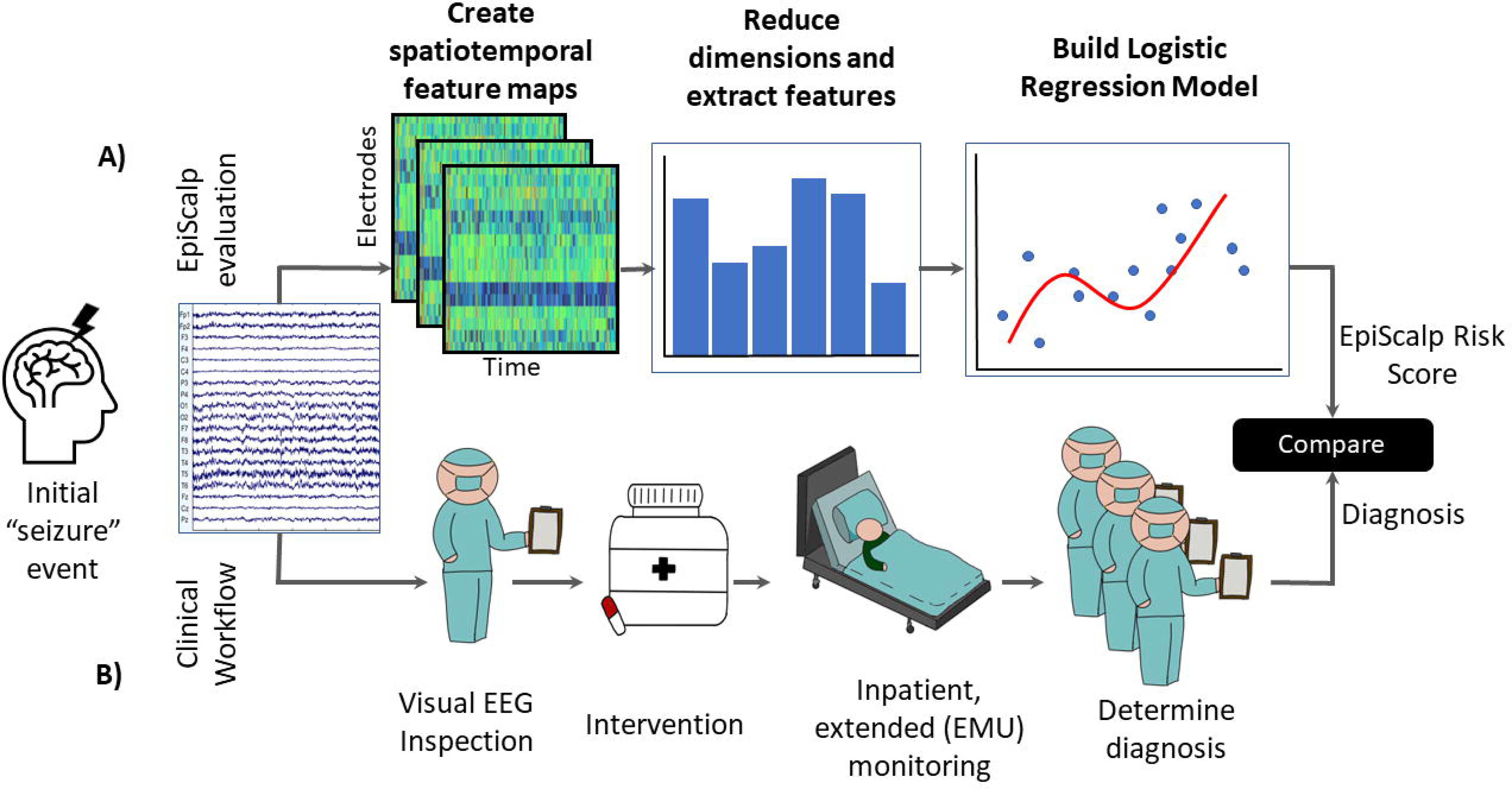

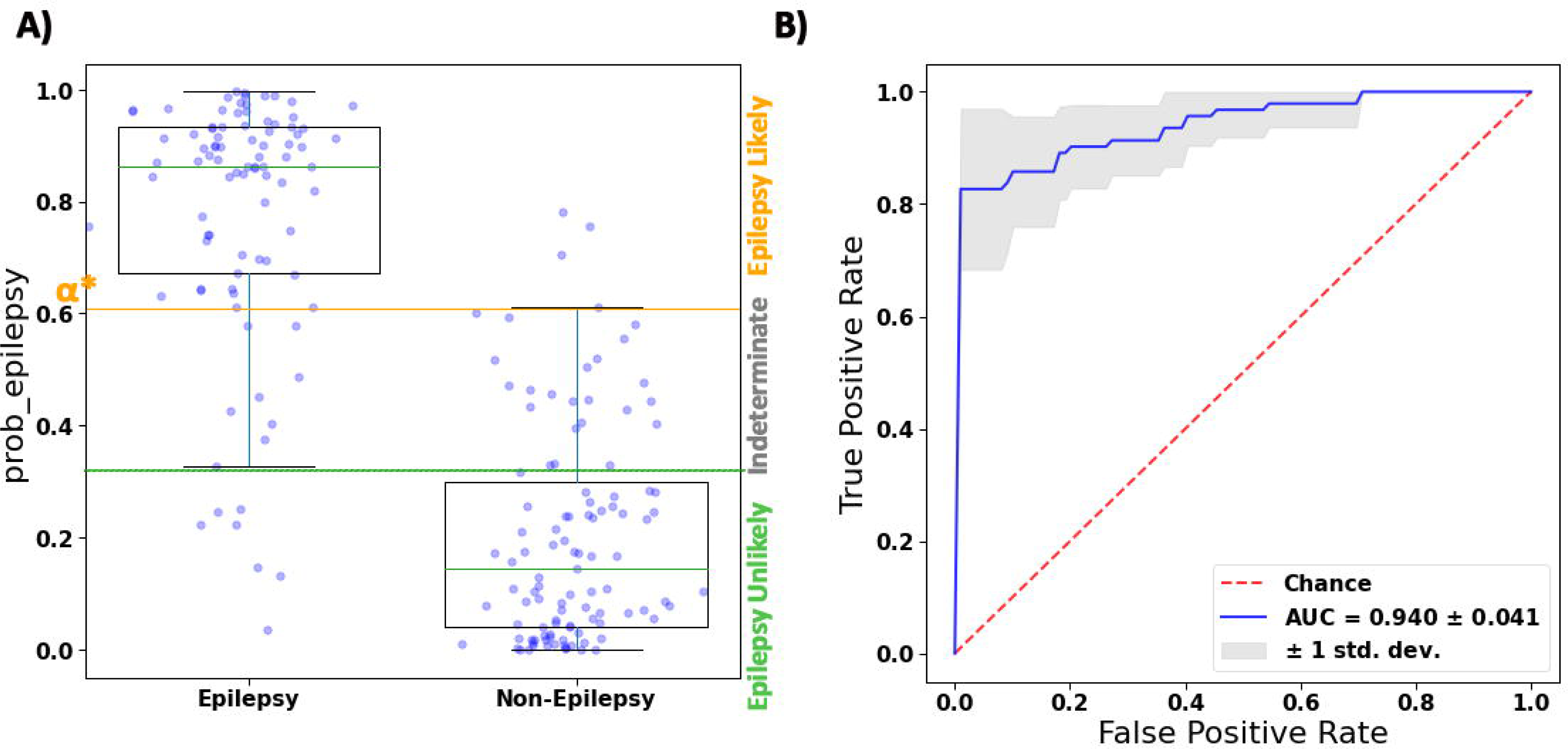

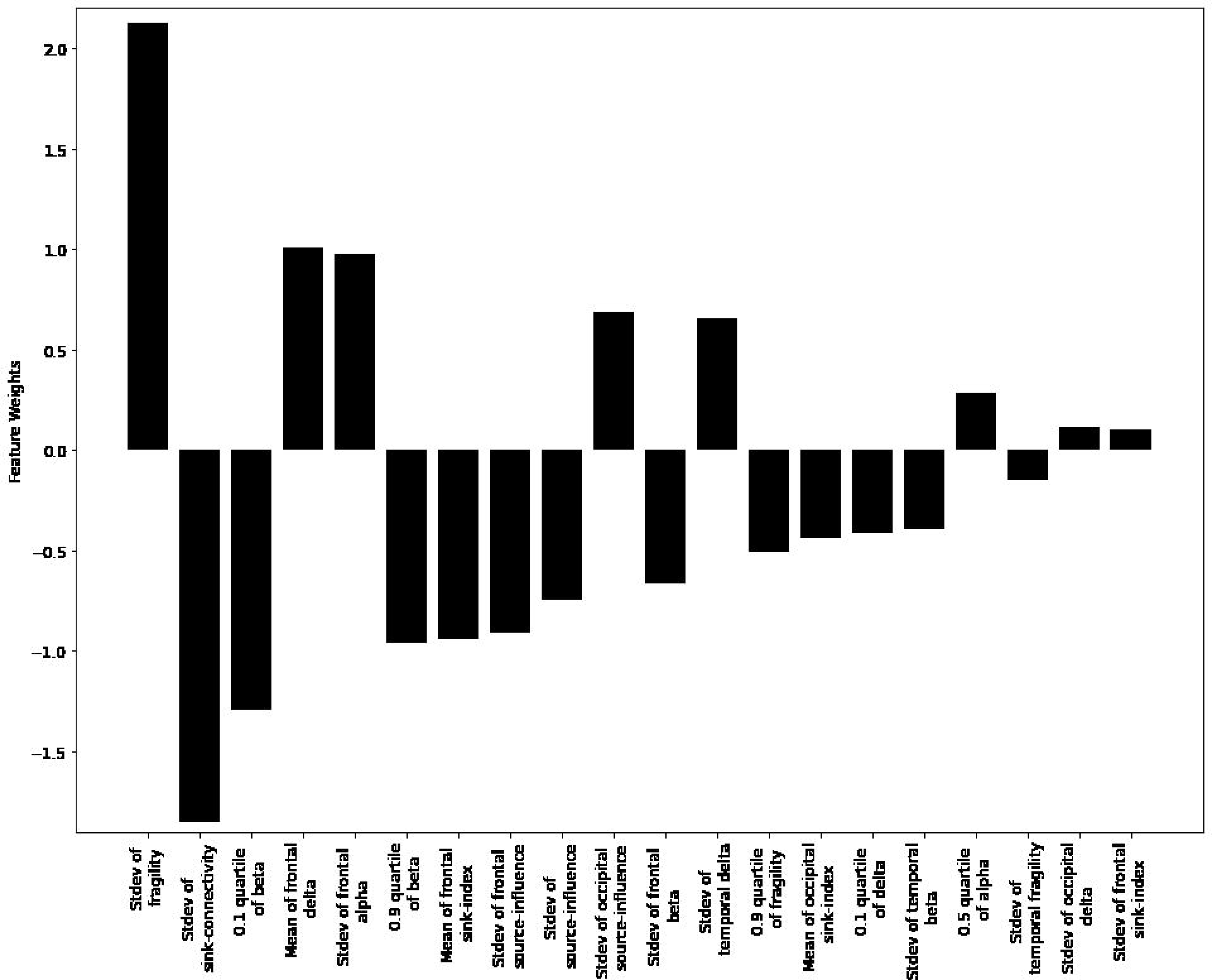

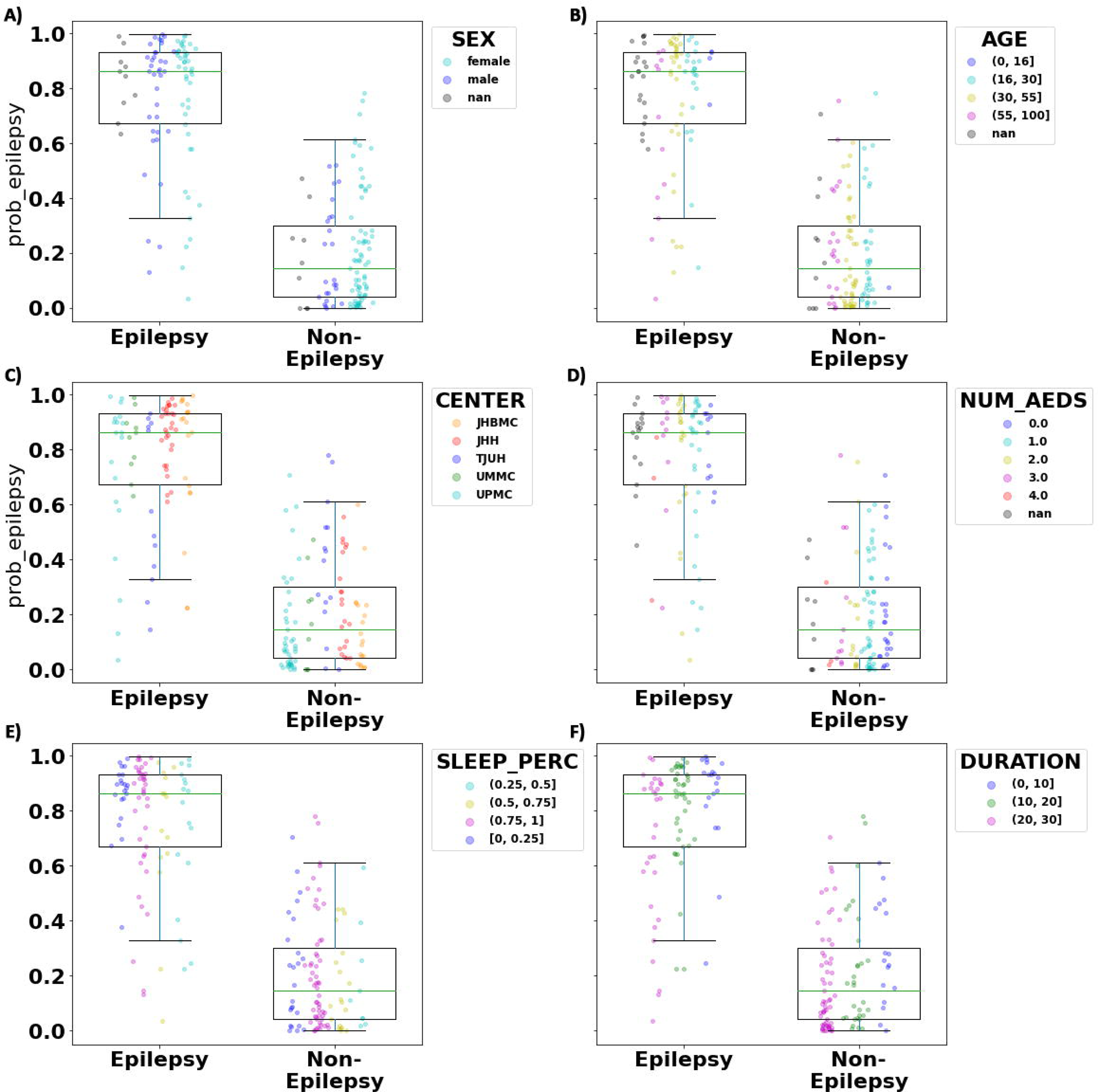

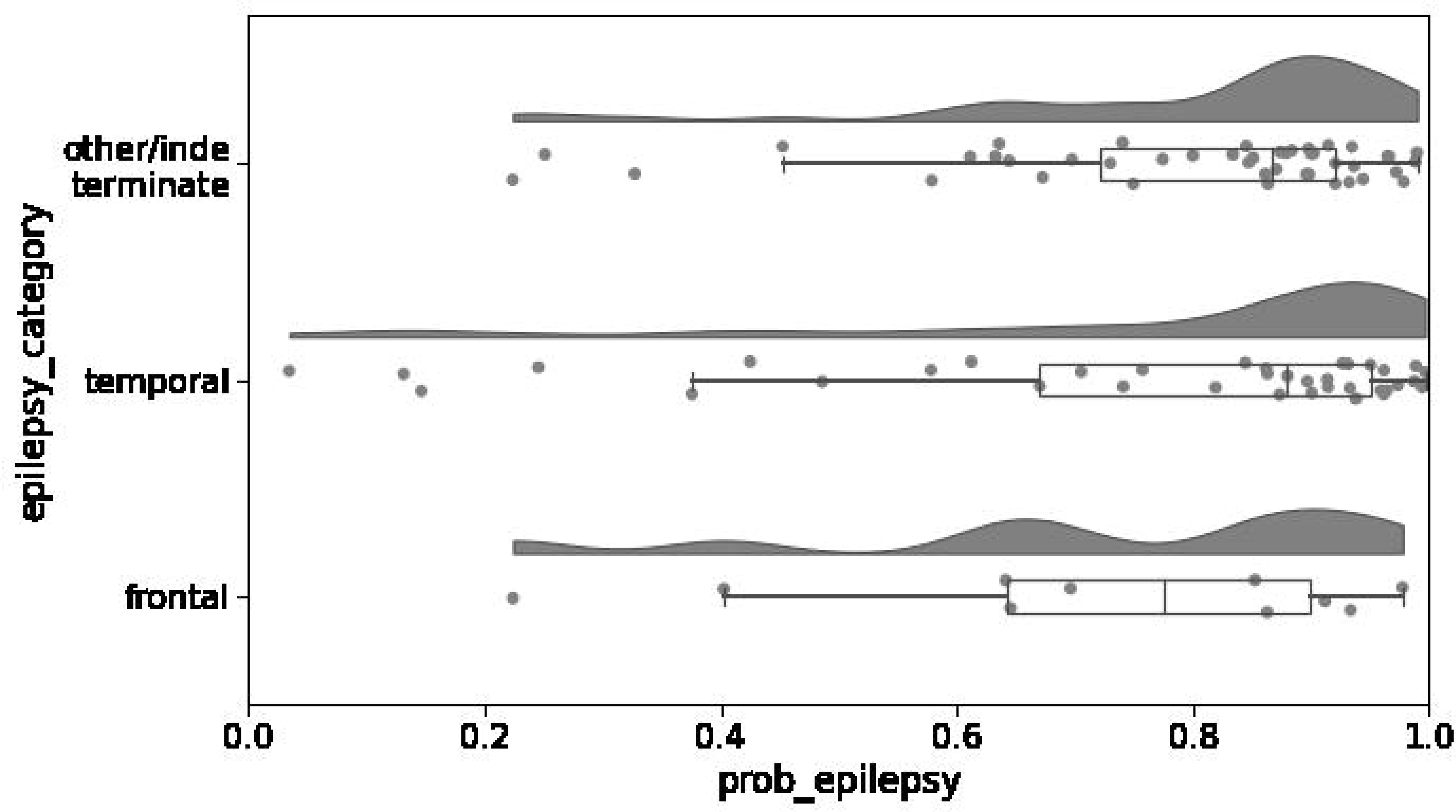

|  | Epilepsy | Non-Epilepsy |
| --- | --- | --- |
| Johns Hopkins Hospital | 50 | 40 |
| Thomas Jefferson University Hospital | 12 | 15 |
| University of Pittsburgh Medical Center | 19 | 43 |
| University of Maryland Medical Center | 10 | 9 |
| Total | 91 | 107 |

| Feature Name | Weight | Metric Type | Dimensionality Reduction | Feature Category |
| --- | --- | --- | --- | --- |
| Stdev of fragility | 2.13 | Fragility | Time-average | Channel (quartile) |
| Stdev of sink-connectivity | -1.85 | Sink-connectivity | weighted time average (PCA-1st component) | Channel (quartile) |
| 0.1 quartile of beta | -1.29 | Spectral analysis - Beta frequency | weighted time average (PCA-2nd component) | Channel (quartile) |
| Mean of frontal delta | 1.01 | Spectral analysis - Delta frequency | weighted time average (PCA-1st component) | Lobes (frontal) |
| Stdev of frontal alpha | 0.98 | Spectral analysis - Alpha frequency | Time-average | Lobes (frontal) |
| 0.9 quartile of beta | -0.96 | Spectral analysis - Beta frequency | weighted time average (PCA-2nd component) | Channel (quartile) |
| Mean of frontal sink-index | -0.94 | Sink-index | weighted time average (PCA-1st component) | Lobes (frontal) |
| Stdev of frontal source-influence | -0.91 | Source-influence | Time-average | Lobes (frontal) |
| Stdev of source-influence | -0.74 | Source-influence | Time-average | Channel (quartile) |
| Stdev of occipital source-influence | 0.69 | Source-influence | weighted time average (PCA-1st component) | Lobes (occipital) |
| Stdev of frontal beta | -0.67 | Spectral analysis - Beta frequency | weighted time average (PCA-2nd component) | Lobes (frontal) |
| Stdev of temporal delta | 0.65 | Spectral analysis - Delta frequency | Time-average | Lobes (temporal) |
| 0.9 quartile of fragility | -0.51 | Fragility | weighted time average (PCA-2nd component) | Channel (quartile) |
| Mean of occipital sink-index | -0.44 | Sink-index | weighted time average (PCA-1st component) | Lobes (occipital) |
| 0.1 quartile of delta | -0.41 | Spectral analysis - Delta frequency | weighted time average (PCA-1st component) | Channel (quartile) |
| Stdev of temporal beta | -0.40 | Spectral analysis - Beta frequency | Time-average | Lobes (temporal) |
| 0.5 quartile of alpha | 0.29 | Spectral analysis - Alpha frequency | weighted time average (PCA-1st component) | Channel (quartile) |
| Stdev of temporal fragility | -0.15 | Fragility | Time-average | Lobes (temporal) |
| Stdev of occipital delta | 0.12 | Spectral analysis - Delta frequency | weighted time average (PCA-1st component) | Lobes (occipital) |
| Stdev of frontal sink-index | 0.10 | Sink-index | weighted time average (PCA-1st component) | Lobes (frontal) |

